# Improved Global Response Outcome After Intradetrusor Injection of Adult Muscle-Derived Cells for the Treatment of Underactive Bladder

**DOI:** 10.1101/2020.06.16.20132779

**Authors:** Jason Gilleran, Ananias C. Diokno, Elijah Ward, Larry Sirls, Deborah Hasenau, Jennifer Giordano, Evelyn Shea, Sarah N. Bartolone, Laura E. Lamb, Michael B. Chancellor

**Author notes:** Corresponding Author: Michael B. Chancellor.

## Abstract

We report on the first regulatory approved clinical trial of a prospective open-label physician-initiated study assessing the safety and efficacy of intradetrusor injected Autologous Muscle Derived Cells (AMDC) treatment for underactive bladder (UAB).

20 non-neurogenic UAB patients were treated. Approximately 50-250 mg of quadriceps femoris muscle was collected using a spirotome 8-gauge needle. The muscles biopsy samples were sent to Cook MyoSite (Pittsburgh, PA) for processing, isolation, and propagation of cells. Research patients received approximately 30 intradetrusor injections of 0.5 mL delivered to the bladder, for a total of 15 mL and 125 million AMDC, performed utilizing a flexible cystoscope under direct vision using topical local anesthesia. Follow-up assessments included adverse events and efficacy via voiding diary and urodynamic testing at 1, 3, 6 & 12-month post-injection. An optional second injection was offered at the end of the 6 months visit.

20 patients received the first injection and all 20 patients requested and received a second injection. Median patient age was 65 years old (range 41-82 years). There were 16 male (80%) and 4 female (20%) patients. Etiology included 7 men (35%) with persistent urinary retention after transurethral resection of the prostate for benign prostatic hyperplasia and 13 patients (65%) with idiopathic chronic urinary retention. At the primary outcome time point of 12 months, 11/19 patients (58%) reported a global response assessment (GRA) ≥ 5, showing slight to marked improvement in their UAB symptoms, compared to 6/20 (30%) patients at 3-months post-injection. No serious procedure or treatment-related adverse events occurred. Noted improvements included: decreased post void residual urine volume, increased voiding efficiency, and decreased catheter use.

Intradetrusor injected AMDC as a treatment for UAB was successfully completed in a 20-patient trial without serious adverse event and with signal of efficacy. Cellular therapy may be a promising novel treatment for catheter dependent chronic urinary retention. A multicenter controlled trial is needed to further assess the promise of regenerative medicine in the treatment of lower urinary tract dysfunction.

## INTRODUCTION

In this study, we evaluated the safety of Autologous Muscle Derived Cells (AMDC) in the treatment of chronic Underactive Bladder (UAB). UAB is caused by deteriorating bladder function with incomplete bladder emptying symptoms including urine frequency, urgency, hesitancy, difficulty starting and/or stopping voiding, incontinence, nocturia, straining to void and recurrent urinary infections [1],[2],[3],[4],[5]. Diokno and associates reported that 22% of men and 11% of women over 60 reported difficulty emptying their bladders [6],[7].

No medications have proven effective in the long-term treatment of UAB, and consequently patients who suffer from UAB are usually managed with clean intermittent self-catheterization (CIC), indwelling urethral or suprapubic catheters, or urinary diversion [8],[1]. Patients using catheters can face several long-term medical difficulties. In one study, 202 long-term indwelling catheter users reported having experienced the following problems: urinary tract infection (31%), blockage of the catheter (24%), leakage 12%, catheter-associated pain (23%), and dislodgement (12%) [9]. The long-term effects of UAB can lead to recurrent infections, bladder and kidney stones, vesicoureteral reflux, and may cause kidney damage [10]. For many individuals, the use of catheters can also be a cause for embarrassment and can negatively impact their work and home life. Moreover, the need for catheterization can have a major impact on quality-of-life (QOL), especially in aging adults. It has been estimated that about 30% of older adults are admitted to long-term care in part due to loss of bladder control [11].

Catheter dependent chronic urinary retention (CUR) is the most severe manifestation of UAB. CUR is defined as an elevated post void residual (PVR) greater than 300 mL that persists for at least 6 months and is documented on 2 or more separate occasions [10].

Regenerative medicine approaches have been reported to treat lower urinary tract dysfunction [2]. AMDC are currently under investigation and have been tested in women with stress urinary incontinence [12],[13],[14]. AMDC have not been associated with serious adverse effects (SAEs) and offer promising efficacy [14]. We hereby report the first trial of using AMDC in the treatment of chronic UAB.

## METHODS

This was a physician sponsored, single center, two-year prospective, open-label, clinical trial, assessing the safety and efficacy of intradetrusor injected AMDC as a treatment for UAB in 20 research participants (ClinicalTrial.gov; NCT02463448). This study received approval from the Beaumont Investigational Review Board (IRB# 2015-134). Written consent from all participants was obtained prior to initiating any study activity. The primary objective of this study was to evaluate the safety of AMDC in the treatment of UAB at 6 months post initial injection. Primary safety endpoints included assessment of AMDC, the biopsy and injection procedures, and post-injection cystoscopy immediately after initial injection and at 6 months post-initial injection.

The secondary objective of the study was to evaluate the safety and efficacy of AMDC in the treatment of UAB at 12 months post-initial injection. Secondary efficacy endpoint measures included Global Response Assessment (GRA) and changes in voiding habits (i.e. frequency, urgency, urine volume voided independently, urine volume voided via catheterization) as recorded on the 3-day bladder diary.

Key inclusion criteria include persons18 years of age or older, history of UAB for at least 6 months with symptoms unresponsive to previous use of medications and/or other treatments, voiding difficulty (complains of difficulty emptying the bladder), post void residual (PVR) ≥ 150 mL, total UAB Questionnaire Score ≥ 3 or 100% reliant on clean intermittent catheterization (CIC) for bladder emptying. Key exclusion criteria include women that are pregnant or planning to become pregnant, bleeding diathesis, anticoagulant therapy, treatment with an investigational device, drug, or procedure for UAB within the last 6 months, history of radiation therapy to the bladder, or pelvic organ prolapse beyond the introitus (e.g., cystocele, rectocele).

### Intervention

All enrolled participants received an initial injection of 125 ⨯ 10^6^ AMDC. If after the 6-month visit the study investigator determined that the subject may benefit from undergoing a second treatment, the patient had the option of receiving another injection of 125⨯ 10^6^ AMDC approximately 10 weeks after the 6-month visit. Therefore, the total possible cell dose delivered over the course of the study was up to 250⨯ 10^6^ AMDC. The AMDC injection was performed via flexible cystoscope. The biopsy and culture technique allowed AMDC to be produced for multiple injections from a single biopsy procedure. Therefore, participants did not have to undergo a second biopsy procedure prior to receiving the second injection.

### Cell Processing Procedure

After obtaining informed consent and undergoing screening activities, study eligibility was determined. Eligible participants returned to the research clinic for a muscle biopsy procedure performed under local anesthesia. AMDC were generated from tissue procured utilizing the Bioncise Spirotome 8 gauge needle from the vastus lateralis. Samples totaling approximately 50-250 mg were collected and sent to Cook MyoSite, Inc. (Pittsburgh, PA) for subsequent culture and expansion. The culture process preferentially expands desirable AMDC, producing a final cell culture that is enriched in myogenic cell content following current Good Manufacturing Practices methodologies. Approximately 12 weeks after the muscle biopsy, the participant returned to the research clinic to receive bladder injections of AMDC. The expanded AMDC was supplied frozen in a cryogenic medium (2ml), and then thawed and diluted with physiological saline (13ml) for injection.

### Injection Procedure

The injection procedure consisted of injections of a total of 125 million AMDC in a total volume of 15ml distributed throughout approximately 30 sites (0.5 ml/injection) throughout the bladder, with the treatment goal of enhancing bladder detrusor contractility. This procedure is similar to intradetrusor injection of botulinum toxin injection. The procedure was performed under local 1% lidocaine anesthesia in a 20 ml intravesical instillation.

### Assessment Measures

Participants completed multiple questionnaires to assess their UAB symptoms and response to AMDC treatment. The Underactive Bladder Questionnaire (UAB-q) and International Consultation on Incontinence Questionnaire Female/Male Lower Urinary Tract Symptoms Modules (ICIQ-F/MLUTS) surveys assess lower urinary tract symptoms and bother. The Global Response Assessment (GRA) measures change in lower urinary tract symptoms after receiving treatment, compared to baseline. The GRA is a 7-point descriptive scale, scores of 1-3 indicate worsening of symptoms, and 5-7 indicate and improvement in symptoms. A response of 4 represents no change in UAB symptoms compared to baseline. These measures were completed by participants at baseline, 3-months, 6-months, and 12-months post-injection.

### Data Analysis

Patient voiding diaries and GRA scores were used to describe treatment efficacy. Outcomes analyzed include volume per voiding event or catheterization, PVR, and voiding efficiency, calculated as

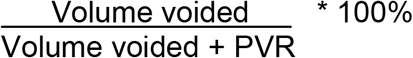

## RESULTS

Twenty non-neurogenic UAB patients were treated. These 20 research participants received the initial AMDC injection, with 20 of the 20 asking for and receiving a second injection. Age of participants ranged from 41-82 years old with a median age of 65 years. There were 16 males (80%) and 4 females (20%). Etiology included 13 patients (65%) with idiopathic chronic urinary retention and seven men (35%) with persistent urinary retention after transurethral resection of the prostate for benign prostatic hyperplasia (**Table 1**). Ten participants were CIC dependent, 9 were mixed voiding (CIC + independent voiding), and 1 was completing voiding independently.

**Table 1.** Patient Demographics.

|  | **Total** | **Male** | **Female** |
| --- | --- | --- | --- |
| **Gender** | 20 | 16 | 4 |
| **Average Age ± SD** | 64.3 ± 11.6 | 62.8 ± 12.1 | 56.8 ± 7.5 |
| **Etiology** |  |  |  |
| **BPH post TURP** | 7 | 7/16 (43.8%) | 0 |
| **Idiopathic** | 13 | 9/16 (56.2%) | 4/4 (100%) |
| **Other Relevant Diseases** |  |  |  |
| **Diabetes** | 1/20 (5%) | 1/16 (6.2%) | 0 |
| **Hx. Pelvic Surgery** | 7/20 (35%) | 4/16 (25%) | 3/4 (75%) |
| **Baseline bladder function** |  |  |  |
| **CIC only** | 10 | 9 | 1 |
| **Mixed void** | 9 | 6 | 3 |
| **Void only** | 1 | 1 | 0 |

Safety was the primary outcome of the study. No AMDC-related adverse were reported. Biopsy-related and injection-related adverse events are listed on **Table 2**. All reported adverse events resolved spontaneously without sequelae. Participants completed the GRA at 3, 6, and 12-months post-injection. As the study progressed, a larger number of participants reported symptom improvement with 11/19 (58%) of patients reporting improvement at 12-months post-injection and 4/19 (21%) reporting no change in symptom quality (**Table 3**). Four patients, of the 20 that received the second injection (21%), have not had their 12-month follow-up visit. One patient reported a mild worsening of symptoms at the 6-month follow-up, but reported no change in symptoms at 12-months.

**Table 2.** Study adverse events.

| **Procedure Related Event** | **Number of patients** |
| --- | --- |
| **Injection Related Events** |  |
| Urinary Tract Infection | 7 |
| Gross Hematuria | 1 |
| Urine Leakage | 1 |
| Rash (from antibiotic) | 1 |
| **Biopsy Related Events** |  |
| Bruised Thigh | 1 |
| Vasovagal Response | 1 |
| Hypoglycemia | 1 |
| Nausea | 1 |
| **Other Event** | **Number of patients** |
| Upper Respiratory Infection | 2 |
| Neck/Shoulder Orthopedic Problems | 2 |
| Atrial Fibrillation | 1 |
| Nonspecific ST-T abnormalities | 1 |

**Table 3.**
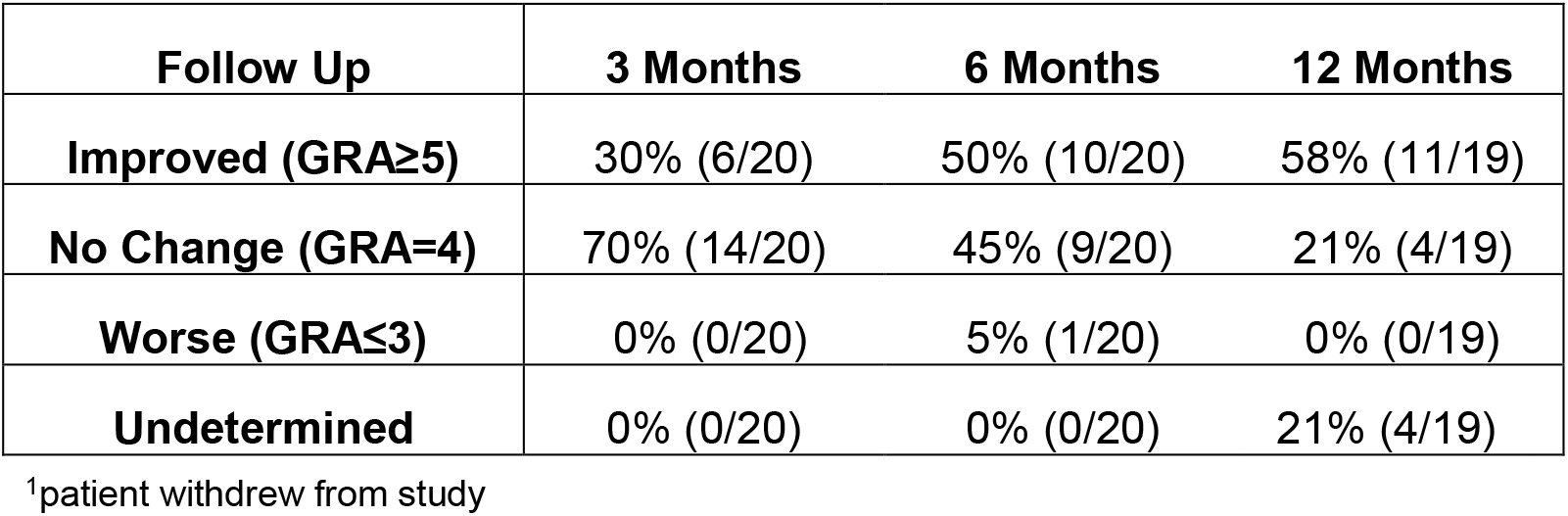
Study Outcomes based on patient-reported GRA.

| **Follow Up** | **3 Months** | **6 Months** | **12 Months** |
| --- | --- | --- | --- |
| **Improved (GRA≥5)** | 30% (6/20) | 50% (10/20) | 58% (11/19) |
| **No Change (GRA=4)** | 70% (14/20) | 45% (9/20) | 21% (4/19) |
| **Worse (GRA≤3)** | 0% (0/20) | 5% (1/20) | 0% (0/19) |
| **Undetermined** | 0% (0/20) | 0% (0/20) | 21% (4/19) |

<sup>1</sup>patient withdrew from study

CIC-dependent patients who reported an improvement in symptom quality (GRA≥5) showed a decrease in urine volume-per-catheterization during their 3-day voiding diaries at each time point, while participants who reported no change (GRA≤4) had no change in their catheterization volume (**Figure 1A**). The same effect can be seen in volume-per-void for mixed-void patients (**Figure 1B**). Participants who improved saw an increase in volume per void, but a decrease in number of voids per day. Participants who reported no improvement saw a decrease in volume per void, with an increase in number of voids per day.

**Figure 1:**
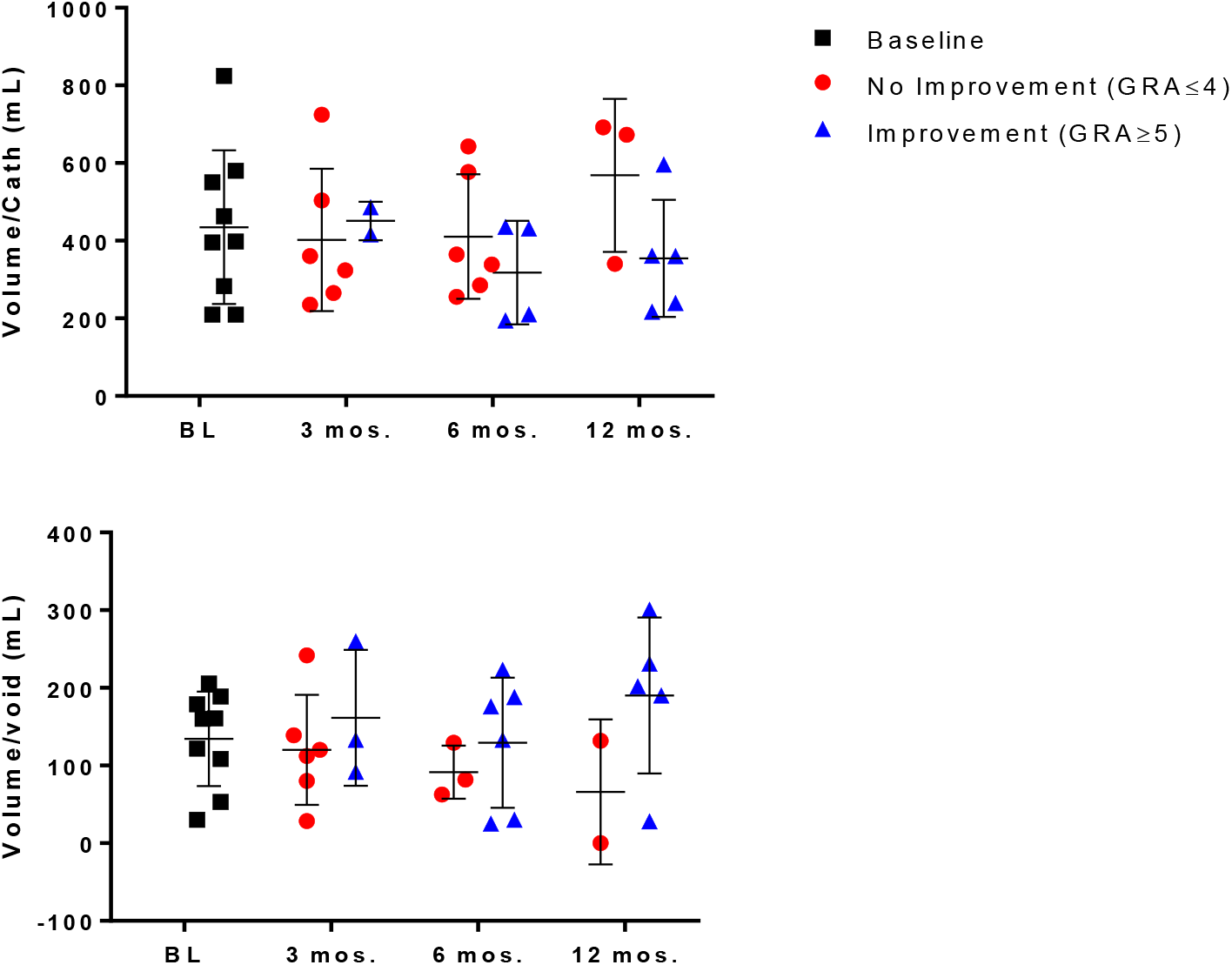
Change in volume/void and volume/catheterization separated by GRA.

PVR of participants with the ability to void was measured at the time of urodynamic testing performed at baseline (n=8), 6-months (n=11), and 12-months post-injection (n=9). Participants showed a general trend in decrease of PVR as the study progressed (**Figure 2A**), with participants who reported symptom improvement (GRA≥5) showing a larger decrease compared to those who reported no change in symptoms (**Figure 2B**).

**Figure. 2:**
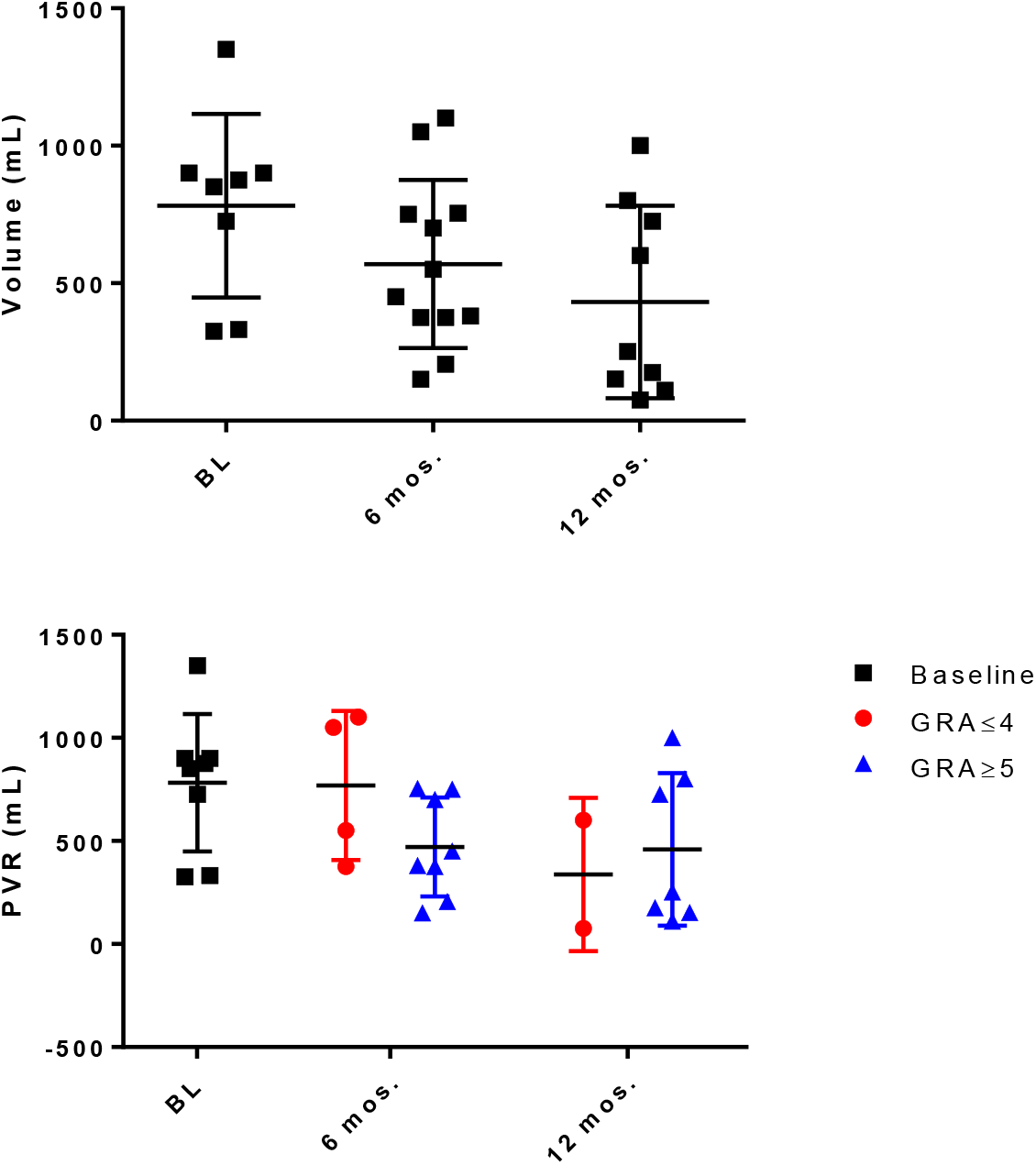
Post void residual volume (y axis in mL) general and separated by GRA.

Voiding efficiency was calculated using participant data for those with measurable void volume and PVR during urodynamic testing. Patients with no measurable void during baseline urodynamics (n=7) were not included in this figure. As the study progressed, average voiding efficiency for all patients increased (**Figure 3**).

**Figure 3:**
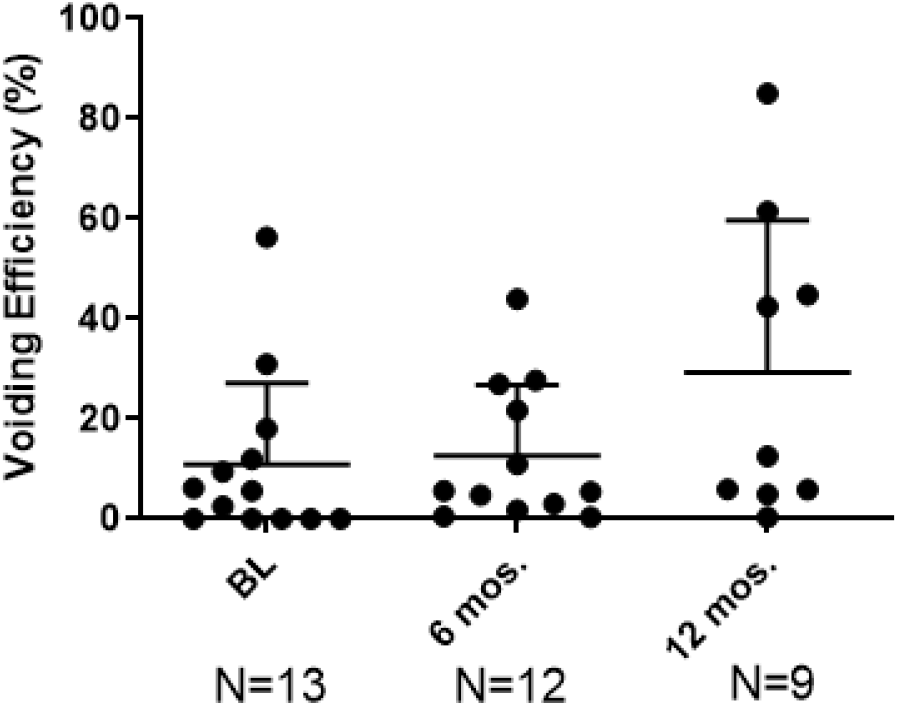
Voiding efficiency.

## DISCUSSION

We report on the first regenerative medicine clinical trial using AMDC for the treatment of UAB with demonstration of procedure safe and encouraging improvement in bladder functioning. In the treatment of female SUI, AMDC is hypothesized to engraft into the urethral muscle layer, lead to formation of new striated muscle, and improve urethral sphincter function. [15],[16],[17]. AMDC do not reabsorb or redistribute, so it may be safe and provide potential long-term outcomes for the treatment of women with SUI. The cystoscope and injection needle are familiar to urologists and urogynecologists and bladder AMDC injection to treat UAB is done in similar fashion as intradetrusor bladder botulinum toxin injection.

The American Urological Association (AUA) proposed a treatment algorithm for CUR in which patients are stratified first by risk and then by symptoms [10]. Based on these features, patients may undergo observation, treatment, or further testing through more invasive urodynamic tests. In asymptomatic patients with existing UAB but without high-risk features like hydronephrosis, stage 3 chronic kidney disease or recurrent UTI, a period of observation before any additional treatment is usually recommended. In symptomatic patients, behavioral management, catheterization, and eventually surgical intervention represent the treatment options.

There are few prospective, randomized, controlled studies supporting conservative behavioral therapy in the UAB population. The use of assisted bladder emptying procedures such as techniques to expel urine (Crede) and voiding by abdominal straining (Valsalva maneuver) create high pressures and are considered potentially hazardous, therefore their use is strongly discouraged by the European Association of Urology (EAU) [18]. According to the EAU guidelines, CIC represents the gold standard for the management of neurogenic lower urinary tract dysfunction. On average, the use of a 12–14 French catheter four to six times per day is needed. Less frequent catheterization results in higher bladder-storage volumes and increased risk of UTI, whereas more frequent catheterization can increase the risk of cross-infection [18], [19]. Although CIC represents the standard of care in bladder management, there are cases where catheterization may not be a suitable method for the management of UAB including the inability to independently catheterize due to poor hand function, lack of a willing caregiver to perform catheterization, abnormal urethral anatomy (false passage), and a low bladder capacity. Furthermore, the inability to self-catheterize is associated with problems that affect QOL, with loss of independence for both the patient and the caregiver. Indwelling catheters, both urethral or suprapubic, are also utilized for short- and long-term management of UAB. Several studies looked at how well indwelling catheters are tolerated compared to CIC and found that a vast majority of patients preferred an indwelling catheter to other type of bladder management for social and practical reasons [20]. However, complications due to bladder stones, decreased bladder capacity and UTI are more common with indwelling catheterization and suprapubic cystostomy compared to other types of bladder management methods [20]. In addition, chronic catheterization has been associated with an increased risk of bladder malignancy, particularly squamous cell carcinoma [21].

Pharmacological therapies for UAB have mainly focused on increasing detrusor smooth muscle contractility, through the enhancement of parasympathetic activity. Bethanechol chloride is a synthetic parasympathomimetic choline carbamate that selectively stimulates muscarinic receptors and that has been available since the 1970s to treat urinary retention [22]. Distigmine bromide is a long-acting anticholinesterase, available for the treatment of postoperative urinary retention and detrusor underactivity. Both these agents have been widely used for UAB management for decades however clinical data show conflicting results about their efficacy. A systemic review of randomized clinical trials concluded that there is little evidence to support the use of muscarinic receptor agonists and/or acetylcholinesterase inhibitors in the treatment of UAB, specifically when adverse effects are taken into account [23],[24].

Sacral neuromodulation (SNM) (Medtronic, MN) was approved over 20 years ago for the indications of inability to completely empty the bladder and the symptoms of OAB. In a registry study from 1993 to 1997, a mixture of OAB and UAB patients was evaluated with SNM. Thirty-one of the 51 UAB patients (61%) were able to eliminate catheter use, and another 16% had a 50% reduction in catheter use [25]. A randomized multicenter trial aimed to evaluate the efficacy of SNM for urinary retention was performed by Jonas et al. [26]. In this study, successful results were initially achieved in 83% of patients who received permanent implant, with 69% of them able to discontinue intermittent catheterization completely. At 18 months, 71% of patients available for follow-up had sustained improvement. SNM is however limited by lack of significant experience in catheter dependent CUR [27]. Myogenic etiology UAB patients are unlikely to respond to nerve stimulation and a percentage of patients who have a successful test stimulation will not improve to the same extent after the permanent implant due to problem with electrode placement.

Other surgical techniques for treating chronic urinary retention associated with neurogenic dysfunction such as nerve re-routing and latissimus dorsi myoplasty have limited long-term data in a highly selected population. Similarly, reductive techniques such as bladder diverticulectomy and reduction cystoplasty still remain the subject of small case reports, therefore the AUA does not recommend these procedures for routine treatment of UAB patients [10].

Patient response to treatment was measured via GRA, diary of voiding and catheterization events and volume and PVR measurement. As the study progressed, a larger number of patients reported at least minimal improvement compared to baseline, with 58% of participants reporting a GRA ≥ 5 at the 12-month follow-up visit. Patients who reported an improvement in their UAB symptoms compared to baseline showed positive changes in volume per voiding event, voiding efficiency, and PVR volume.

There were no significant changes on urodynamic parameters. These results suggest that AMDC-UAB injections could be a promising treatment option for UAB patients with CUR.

The limitations of this study include small sample size and lack of randomization, but this is in-line with a first-in-human, physician initiated, feasibility study of safety. UAB is one of the greatest areas of unmet need and yet has the greatest opportunity for advancement in functional urology. The disease is common, severe, and lacks effective treatment. There is great international interest in the research of UAB and we believe the path forward should include engaging regulatory agencies around the world that can harmonize and formalize guidance for regulatory trial designs for therapeutics for UAB.

## Conclusion

Intradetrusor injected AMDC for the treatment of UAB was successfully completed in a 20-patient trial without any serious adverse events. Improvement, as indicated by the GRA, decreased PVR, and improvement of voiding efficiency was seen in many subjects who were catheter dependent at baseline. Cellular therapy may be a promising novel treatment for catheter dependent chronic urinary retention. Multicenter controlled trials are needed to further assess the promise of regenerative medicine in the treatment of lower urinary tract dysfunction.

## Data Availability

Data will be made available upon request.

## Research Involving Human Participants

All procedures performed in studies involving human participants were in accordance with the ethical standards of the institutional and/or national research committee and with the 1964 Helsinki declaration and its later amendments or comparable ethical standards.

## Informed Consent

Informed consent was obtained from each subject before performing any screening procedures by the Principal Investigator, co-Investigator, or research coordinator.

## Funding and Acknowledgement

The Aikens Research Center at Beaumont Health System (Royal Oak, MI, USA). The authors would like to acknowledge the NIA, NIDDK, the Underactive Bladder Foundation (www.underactivebladder.org).

## Legends

**Figure 1. Impact of AMDC treatment on urinary output over time by GRA subgroup**. Average urinary output was calculated from a 3-day bladder diary given at the indicated time point post AMDC treatment. Patients were divided into subgroups based on their perception of treatment impact on their symptoms as measured by GRA. GRA scores above 4 reported an improvement in symptoms compared to baseline; GRA scores of 4 reported no improvement in symptoms compared to baseline; GRA scores less than 4 reported worse symptoms compared to baseline. The number of patients in the GRA≥5 group increased over time, independent of catheterization dependent or mixed voiding, showing treatment efficacy. **1A)** Catheter-dependent patients who reported a GRA≥5 showed a decrease in volume per catheterization, while patients who reported a GRA≤4 showed an increase in volume. **1B)** In mixed voiding patients, those who reported a GRA≥5 showed an increase in volume per event, while patients who reported a GRA≤4 exhibited a decrease in volume per event. Volume/cath (mL)= volume per catheterization; BL=baseline measurement at study enrollment; mos=month.

**Figure 2. Impact of AMDC treatment on Post-Void Residual volume over time**. Average PVR volume was calculated from urodynamics testing performed at follow-up visits. **1A)** For patients with measurable volumes, average PVR decreased from baseline to 12-months post-injection. **1B)** When separated by GRA, patients who reported a GRA≥5 showed a larger decrease in PVR volume at 6-months post-injection. The number of patients in the GRA≥5 group increased over time, independent of catheterization dependent or mixed voiding, showing treatment efficacy. BL=baseline measurement at study enrollment; mos=month.

**Figure 3. AMDC treatments causes increase in voiding efficiency over time**. Voiding efficiency was calculated from urodynamics testing performed at baseline and follow-up visits. 7 patients were not able to void during baseline urodynamic testing. For patients with measurable voiding volume, voiding efficiency increases over time, showing AMDC treatment efficacy. BL=baseline measurement at study enrollment; mos=month.

